# Healthcare Costs and Healthcare Utilization Outcomes of Vitamin D3 Supplementation at 5000 IU Daily During a 10.9 month Observation Period within a Pragmatic Randomized Clinical Trial

**DOI:** 10.1101/2023.09.25.23296104

**Authors:** Patrick J. LaRiccia, Teresa Cafaro, Dibato John, Noud van Helmond, Ludmil V. Mitrev, Brigid Bandomer, Tracy L. Brobyn, Krystal Hunter, Satyajeet Roy, Kevin Q. Ng, Helen Goldstein, Alan Tsai, Denise Thwing, Mary Ann Maag, Myung K. Chung

## Abstract

Vitamin D insufficiency has been linked to multiple conditions including bone disease, respiratory disease, cardiovascular disease, diabetes, and cancer. Observational studies indicate lower healthcare costs and healthcare utilization with sufficient vitamin D levels. The secondary aims of our previously published pragmatic clinical trial of vitamin D3 supplementation were comparisons of healthcare costs and healthcare utilization. Comparisons were made between the vitamin D3 at 5000 IU supplementation group and a non-supplemented control group. Costs of care between the groups were not statistically different. Vitamin D3 supplementation reduced healthcare utilization in four major categories: hospitalizations for any reason (rate difference: -0.19 per 1000 person-days, 95%-CI: -0.21 to -0.17 per 1000 person-days, *p* < 0.0001); ICU admissions for any reason (rate difference: -0.06 per 1000 person-days, 95%-CI: -0.08 to -0.04 per 1000 person-days, *p* < 0.0001); emergency room visits for any reason (rate difference: -0.26 per 1000 person-days, 95%-CI: -0.46 to -0.05 per 1000 person-days, *p* = 0.0131; and hospitalizations due to Covid-19 (rate difference: -8.47X10^−3^ per 1000 person-days, 95%-CI: -0.02 to -1.05X10^−3^ per 1000 person-days, *p* = 0.0253). Appropriately powered studies of longer duration are recommended for replication of these utilization findings and analysis of cost differences.

## 1. Introduction

Vitamin D insufficiency has been linked to multiple conditions including bone disease [1], respiratory disease [2-4], cardiovascular disease [5], diabetes [6], and cancer [7]. Low vitamin D levels are also associated with in-hospital mortality [8], obesity [9], older age [9], and non-Hispanic Black race [9]. Vitamin D deficiency was closely linked to increased healthcare costs and healthcare utilization in veterans in Northeastern Tennessee: overall cost, emergency room visits, clinic visits, inpatient services, and hospital stays were greater in the vitamin D deficient patients when compared to those with adequate levels [10]. Similarly, vitamin D deficiency was associated with increased healthcare costs at six Veterans Affairs Medical Centers in the Southeastern United States [11,12]. In a community hospital study of 258 patients admitted to the surgical intensive care unit (ICU), vitamin D deficiency was associated with increased cost, length of stay, and mortality [13]. In another community hospital study of 565 patients, patients with vitamin D levels less than 18 ng/ml had higher hospital ward costs and higher ICU costs. They also had more frequent myocardial infarctions, ventilator-associated pneumonias, and longer hospital ward and ICU stays [14]. In two German independent population-based studies (n=7217 total), vitamin D deficiency was associated with increased total annual costs, outpatient costs, inpatient costs, and hospital stays [15]. Based on these studies, we postulated that people with sufficient vitamin D levels have lower healthcare costs and less healthcare utilization than those with lower vitamin D levels.

A previously published pragmatic trial by our group demonstrated that vitamin D3 supplementation at 5000 IU/day reduces influenza-like illness in hospital workers [2]. Herein we describe the results of the secondary aims of this study which were to assess healthcare costs and healthcare utilization outcomes [2].

## 2. Materials and Methods

Details of our pragmatic randomized clinical trial examining the effects of daily intake of 5000 IU of vitamin D3 on the incidence of influenza-like illness in healthcare workers have been published earlier [2]. Included in that publication are the CONSORT flow diagram, CONSORT checklist, and Clinical Trial Registration number). The local Institutional Review Board approved the study (IRB #20-455). Here we describe our methods of data acquisition and analysis of our secondary aims of comparing healthcare costs and healthcare utilization in the control and intervention groups.

### 2.1. Subjects

Subjects were employees of an inner-city university hospital who were at least 18 years of age. All subjects analyzed in the costs and utilization part of the study were insured for their healthcare through the university hospital. Subjects who were not insured through the university hospital healthcare plan were not included in the analyses as their healthcare costs and utilization records were not available. In addition subjects in the passive control group were those who voluntarily completed a survey that included their informed consent, demographics, and medical history which were used for comparison with the intervention group’s demographics and clinical characteristic.

### 2.2. Intervention and control groups

As indicated above the intervention group received 5000 IU of vitamin D3 per day for 9 months. The passive control group received no specific instructions and were followed from the start of the study until the last participant of the intervention group completed 9 months of vitamin D3 supplementation.

### 2.3. Observation Periods

The observation periods were the sum of individually calculated, de-identified subject data for each group. The individual intervention subjects’ observation period began on their date of first dose (plus sixty days) or the date their insurance coverage began, whichever was later; their observation period ended on the date of their last dose or the date their insurance coverage was terminated, whichever was earlier.

The individual control subjects’ observation periods began on the date of the first *intervention* subject’s first dose (plus sixty days) or the date insurance coverage began for the control subject, whichever was later; it ended on the date of the last *intervention* subject’s last dose or the date insurance coverage was terminated for the control subject, whichever was earlier.

Sixty days was added to the date of first dose of vitamin D3 for the intervention group subjects as this is the time period known to achieve therapeutic vitamin D blood levels [16]. The first intervention subject’s first dose plus sixty days was January 2, 2021. The last intervention subject’s last dose was November 23, 2021. The overall time span observed for all subjects combined was 10.9 months (326 days). The person-time denominators for the control and intervention groups were 590,348 and 37,935 person-days, respectively.

### 2.3. Data Acquisition

De-identified data on healthcare costs and healthcare utilization was obtained from the administrators of the university hospital employee insurance plan.

### 2.4. Measurements and Statistical Analysis

#### 2.4.1. Demographics and Clinical Characteristics

Demographics and comorbidity data were collected from both groups via survey. All subjects in the passive control group were invited to voluntarily complete a survey that included their informed consent, demographics, and medical history which were used for comparison with the intervention group’s demographics and clinical characteristic.

Descriptive statistics were used for describing the demographics and comorbidities of the intervention and control groups. To provide an objective means to identify meaningful differences in demographic and clinical characteristics between the intervention and control groups, we used standardized mean differences with a cutoff of 20% or 0.20 [17-19]. Costs and utilization data were available only on subjects who were insured by the university hospital.

#### 2.4.2. Healthcare Costs

Healthcare costs for the control and intervention groups were determined for six categories including total billed charges for any reason; cost of hospitalizations due to COVID-19; cost of ICU admissions due to COVID-19; cost of ventilator use due to COVID-19; medical pharmacy prescription costs for any reason; and freestanding prescription costs for any reason. All costs were determined by the billed charges for each category. The mean cost per person-day (standardized mean) was calculated for each category. Differences in standardized means between control and intervention groups were assessed using Wilcoxon rank-sum tests to determine statistical significance. The alpha level was set at 0.05. All analyses were done using SAS 9.4 (SAS Institute, Inc., Cary, NC).

#### 2.4.3. Healthcare Utilization

Healthcare utilization was determined for fifteen categories: 1) number of hospitalizations for any reason; 2) number of ICU admissions for any reason; 3) number of emergency room visits for any reason; 4) number of hospitalizations due to Covid-19; 5) number of ICU admissions due to Covid-19; 6) all other outpatient units for any reason; 7) number of urgent care visits for any reason; 8) number of primary care physician units for any reason; 9) number of nurse practitioner units for any reason; 10) all other professional units for any reason; 11) number of medical pharmacy units for any reason; 12) number of freestanding prescriptions for any reason; 13) number of ventilator use for any reason; 14) number of ventilator use due to Covid-19; and 15) number of deaths for any reason.

Incidence rates (number of events per person-days) for all utilization categories were calculated and compared between control and intervention groups using count models (Poisson (P), negative binomial (NB), and zero inflated negative binomial (ZINB)) with person-days used at offset. The model with the smallest Akaike Information Criterion (AIC) and Bayesian Information Criterion (BIC) values was chosen for each event. All analyses were done using SAS 9.4 (SAS Institute, Inc., Cary, NC) and conclusions made at 5% significance level.

## 3. Results

### 3.1 Subjects

The sample size for the intervention group and passive control group was 196 and 1,958, respectively, as this was the number study subjects who were insured by the university hospital.

### 3.2. Demographics and Clinical Characteristics

Demographics and clinical characteristics were similar in the control and intervention groups, Table 1. We compared intervention group subjects (196) to control group subjects who voluntarily provided their demographic and comorbidity data via survey (444 out of 1,958). We found no relevant differences between the groups for a range of demographic and clinical characteristics, except for age, Hispanic or Latino ethnicity, and Not Hispanic or Latino ethnicity, each of which was slightly above the predefined standardized difference threshold of 0.20 (the standardized difference was 0.23 in each case).

**Table 1.** Demographic and clinical characteristics of the vitamin D supplementation and control groups.

|  | Vitamin D3<br>(n = 196) | Control<br>(n=444) | Standardized<br>Difference |
| --- | --- | --- | --- |
| Age at enrollment in years, mean ± SD | 47 ± 12 | 50 ± 13 | 0.23 |
| **Gender, n (%)** |  |  |  |
| Man | 46 (23) | 106 (24) | 0.01 |
| Woman | 149 (76) | 337 (76) | 0 |
| Other | 1 (0.5) | 1 (0.2) | 0.05 |
| **Race, n (%)** |  |  |  |
| American Indian / Alaska Native | 1 (0.5) | 1 (0.2) | 0.05 |
| Asian | 9 (5) | 26 (6) | 0.06 |
| Black/African American | 23 (12) | 41 (9) | 0.08 |
| Native Hawaiian/other Pacific Islander | 2 (1) | 0 (0) | 0.14 |
| White | 144 (73) | 348 (78) | 0.12 |
| More than one race | 6 (3) | 14 (3) | 0.01 |
| Other | 11 (6) | 14 (3) | 0.12 |
| **Ethnicity, n (%)** |  |  |  |
| Hispanic or Latino | 22 (11) | 22 (5) | 0.23 |
| Not Hispanic or Latino | 174 (89) | 419 (95) | 0.23 |
| **Body mass index** in kg/m<sup>2</sup>, mean ± SD | 30 ± 6 | 29 ± 7 | 0.17 |
| **Comorbidities, n (%)** |  |  |  |
| Cardiovascular disease | 48 (24) | 130 (29) | 0.11 |
| Respiratory disease | 32 (16) | 84 (19) | 0.07 |
| Eye disease | 9 (5) | 14 (3) | 0.07 |
| Gastrointestinal disease | 79 (40) | 169 (38) | 0.05 |
| Urological disease | 14 (7) | 52 (12) | 0.16 |
| Liver disease | 3 (2) | 6 (1) | 0.02 |
| Hematological disease | 23 (12) | 40 (9) | 0.09 |
| Dermatological disease | 35 (18) | 57 (13) | 0.14 |
| Diabetes | 13 (7) | 36 (8) | 0.06 |
| Endocrine disease (other) | 28 (14) | 58 (13) | 0.04 |
| Malignant disease | 11 (6) | 24 (5) | 0.01 |
| History of vitamin D deficiency, n (%) | 47 (24) | 137 (31) | 0.15 |
| Previous COVID-19, n (%) | 12 (6) | 19 (4) | 0.08 |

### 3.3. Healthcare Costs

The total billed costs in the control group was $41,109,649.83 while the total billed costs in the intervention group was $2,318,500.31. The person-time denominators for the control and intervention groups were 590,348 and 37,935 person-days, respectively.

Three of the six measured parameters indicated less costs in the intervention group. Two parameters indicated less costs in the control group. One parameter indicated no difference at all. There were no statistical differences in any of the cost comparisons. There was a statistical trend in the free-standing pharmacy cost comparison indicating less cost for the control group. See Table 2.

**Table 2.** Standardized Costs by Treatment Groups in US Dollars.

|  | Control (N= 1,958) |  | Intervention (N= 196) |  | Differ-<br>ence | 95%-CI | p-Value |
| --- | --- | --- | --- | --- | --- | --- | --- |
|  | Mean<br>(SD) | Median<br>(Q1, Q3) | Mean<br>(SD) | Median<br>(Q1, Q3) |  |  |  |
| Total billed charges for any reason | 69.3 (179) | 20.1 (6.7, 61.9) | 61.3 (103) | 22.2 (8.5, 74.1) | -8.04 | -24.5 to 8.4 | 0.36 |
| Cost of hospitalizations due to Covid-19 | 0.56 (12.5) | 0 (0, 0) | 0 (0) | 0 (0, 0) | -0.56 | -2.3 to 1.2 | 0.48 |
| Cost of ICU admissions due to Covid-19 | 0.33 (8.9) | 0 (0, 0) | 0 (0) | 0 (0, 0) | -0.33 | -0.72 to 0.06 | 0.58 |
| Cost of ventilator use due to Covid-19 (zeros entry) |  |  |  |  |  |  |  |
| Medical pharmacy prescription costs for any reason | 6.4 (92.4) | 0 (0, 0) | 7.4 (51.4) | 0 (0, 0) | 1.05 | -7.3 to 9.4 | 0.52 |
| Freestanding prescription costs for any reason | 9.05 (38.3) | 1.4 (0.2, 5.4) | 13.4 (34.9) | 1.6 (0.4, 8.1) | 4.4 | -1.2 to 9.9 | 0.07 |

SD-Standard Deviation; CI-Confidence Interval. Difference uses control group as reference.

### 3.4. Healthcare Utilization

Four of the 15 measured parameters comparing the control group with the intervention group showed a statistically significant difference indicating lower healthcare utilization in the intervention group. The four parameters were: number of hospitalizations for any reason; number of ICU admissions for any reason; number of emergency room visits for any reason; and number of hospitalizations due to Covid-19.

There was a trend toward statistical significance for the number of urgent care visits for any reason and the number of ICU admissions due to Covid-19, indicating fewer in the intervention group. Five parameters indicated greater utilization in the intervention group but were not statistically significant: all other outpatient units for any reason, number of primary care physician units for any reason, all other professional units for any reason, number of medical pharmacy units for any reason, and number of freestanding prescriptions for any reason. One of the four remaining comparisons, number of nurse practitioner units for any reason, showed decreased utilization in the intervention group without statistical significance. See Table 3. The last three comparisons, number of ventilator use for any reason, number of ventilator use due to Covid-19, and number of deaths for any reason, showed no difference at all (all entries were zero thus are not listed in Table 3).

**Table 3.**
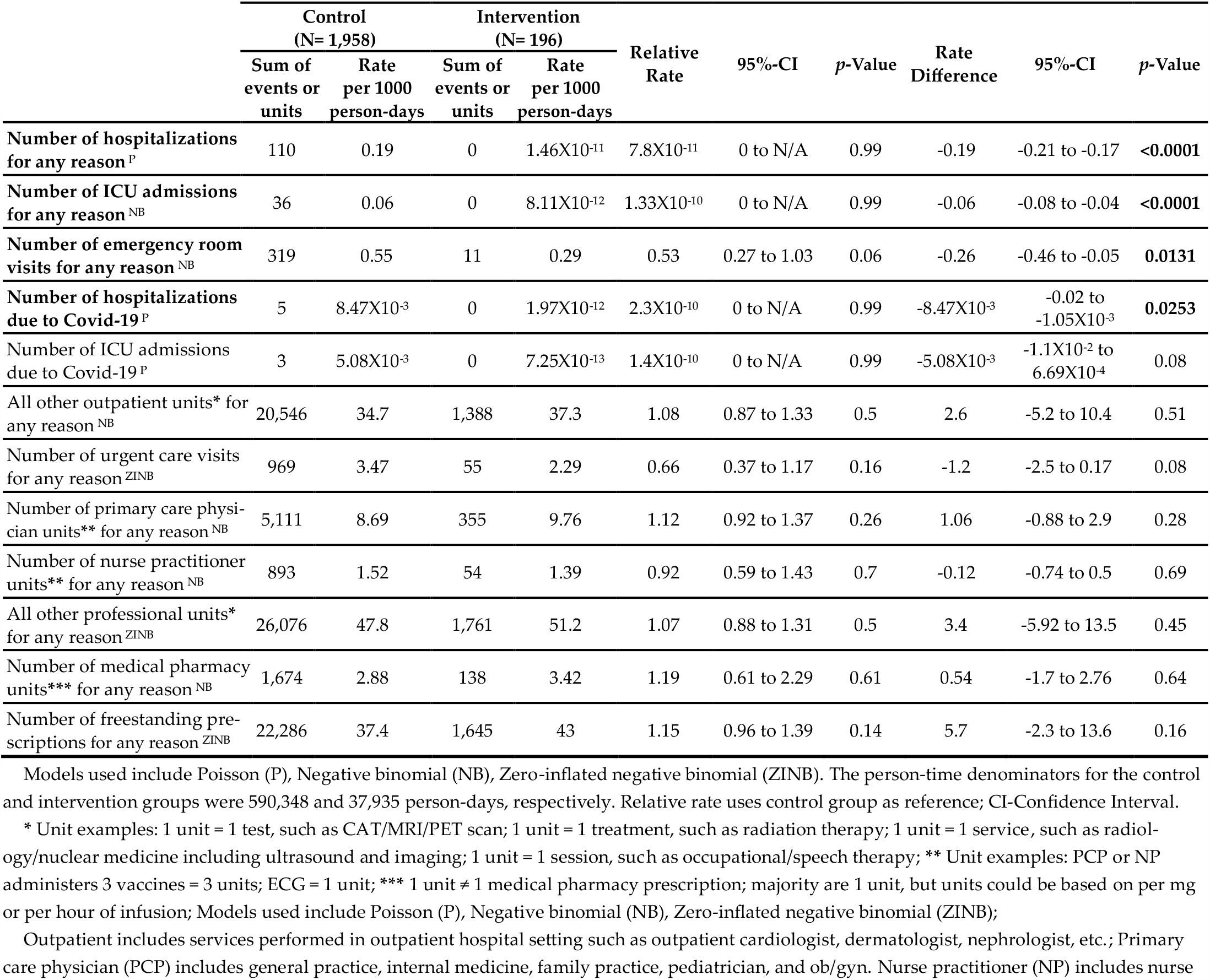
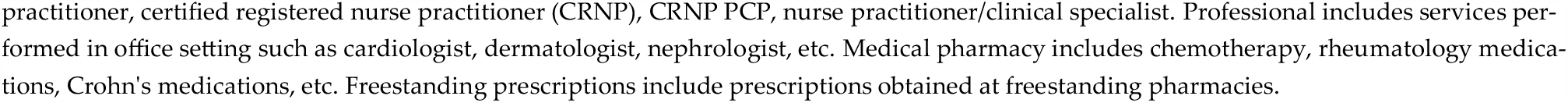
Comparisons of Utilization Between Control and Intervention Groups.

## 4. Discussion

### 4.1. Principal Findings

The insurance claims data of subjects randomized to the vitamin D3 intervention arm and of those subjects randomized to the passive control group in a pragmatic randomized clinical trial was examined for healthcare costs and healthcare utilization. Six healthcare cost parameters and 15 healthcare utilization parameters were evaluated. In the vitamin D3 intervention group there were non-statistically significant decreases in: total billed charges for any reason; cost of hospitalization due to Covid-19; and cost of ICU admissions due to Covid-19. Utilization claims data indicated four areas in which the intervention group showed statistically significant decreases in healthcare utilization: number of hospitalizations for any reason; number of ICU admissions for any reason; number of emergency room visits for any reason; and number of hospitalizations due to Covid-19.

The healthcare utilization results of this study that took place in an East Coast inner-city university hospital in the United States concur with the studies that were performed in Veterans Administration (VA) Medical Centers in Northeastern Tennessee, VA Medical Centers in the Southeastern US, two US community hospitals, and two independent population studies in Germany [10-15] as mentioned above. However, while directional trends in costs were similar in the present study, there were no statistically significant cost differences. A total sample size of 6,098 would have been required to provide sufficient power to detect a mean difference of $8.04/person-day. Our total sample size (both groups combined) was 2,154. Had our study been longer, it is possible that there would have been greater cost differentials. The above studies were cross-sectional and retrospective. Our study was a prospective pragmatic clinical trial, which when combined with the other studies, indicates that despite differing populations, geographic locations, and methodology there is a degree of convergence toward healthcare utilization reduction in the vitamin D3 sufficient groups.

### 4.2. Methodological Considerations

A limitation of this study was that complete data was not available for either the intervention or the control group, as claims data was only obtainable for those subjects who were insured by the hospital healthcare plan. The two groups insured by this plan demonstrated similar demographic and co-morbidity characteristics.

Future research using claims data can be useful in confirming or refuting that daily vitamin D3 intake at 5000 IU can reduce healthcare costs and utilization. Such studies may be particularly helpful when conducted in the context of large employment entities such as university healthcare systems, large corporations, health insurance companies, and health maintenance organizations, to ensure generalizability of results. Quasi-experimental designs could be utilized where a cohort’s baseline healthcare costs and utilization are compared to a prospective interval during which vitamin D3 is given. Randomized placebo-controlled trials can be ethically problematic as observational studies indicate some groups to be more vulnerable to diseases associated with lower vitamin D levels [1,4-9]. Another possibility is a large cohort study with propensity score matching. Future research also needs to consider other micronutrients that may potentiate the benefit from vitamin D3 such as magnesium [20,21].

## 5 Conclusions

In conclusion, 5000 IU of vitamin D3 taken daily reduced hospitalizations for any reason, emergency room visits for any reason, ICU admissions for any reason and hospitalizations due to Covid-19 over a 10.9 month time span. Adequately powered studies of longer duration are recommended.

## Supplementary Materials

None

## Author Contributions

Conceptualization, Patrick J. LaRiccia, Noud van Helmond and Myung Chung; Data curation, Teresa Cafaro; Formal analysis, Patrick J. LaRiccia, Teresa Cafaro, Dibato John and Noud van Helmond; Funding acquisition, Noud van Helmond; Investigation, Patrick J. LaRiccia, Teresa Cafaro, Noud van Helmond and Ludmil Mitrev; Methodology, Patrick J. LaRiccia, Dibato John and Noud van Helmond; Project administration, Patrick J. LaRiccia, Teresa Cafaro and Noud van Helmond; Resources, Patrick J. LaRiccia and Teresa Cafaro; Software, Teresa Cafaro and Dibato John; Supervision, Patrick J. LaRiccia, Noud van Helmond and Ludmil Mitrev; Validation, Patrick J. LaRiccia, Teresa Cafaro and Noud van Helmond; Visualization, Patrick J. LaRiccia, Teresa Cafaro, Noud van Helmond and Ludmil Mitrev; Writing – original draft, Patrick J. LaRiccia; Writing – review & editing, Patrick J. LaRiccia, Teresa Cafaro, Dibato John, Noud van Helmond, Ludmil Mitrev, Brigid Bandomer, Tracy Brobyn, Krystal Hunter, Satyajeet Roy, Kevin Ng, Helen Goldstein, Alan Tsai, Denise Thwing, Mary Ann Maag and Myung Chung (review only). All authors have read and agreed to the published version of the manuscript.

## Institutional Review Board Statement

The study was conducted in accordance with the Declaration of Helsinki and approved by the Institutional Review Board of Cooper University Hospital (protocol code 20-455 and date of approval 20 August 2020).

## Informed Consent Statement

Informed consent was obtained from all subjects involved in the study.

## Data Availability Statement

The data presented in this study are available on request from the corresponding author. The data are not publicly available due to potential privacy concerns.

## Acknowledgments

We extend gratitude to Susan J. Lamon, Jennifer A. Tracy, Justin Frisby, Nicholas Torney Sr., Jermaine Parker, Independence Administrators (Elissa Marsicano, Sheila Aber, Aaron Smith-McLallen, Sonia Cordner and James Kelly), and to the healthcare workers who participated in this study.

